# Total NT-proBNP, a novel biomarker in atrial fibrillation. A mechanistic analysis of the GISSI-AF trial

**DOI:** 10.1101/2020.11.10.20222349

**Authors:** Lidia Staszewsky, Jennifer Meessen, Deborah Novelli, Ulla Wienhues-Thelen, Marcello Disertori, Aldo P Maggioni, Serge Masson, Gianni Tognoni, Maria Grazia Franzosi, Donata Lucci, Roberto Latini, GISSI-AF Investigators.

## Abstract

**Objective:** (1) to test the association with prevalent and incident atrial fibrillation (AF), and prognosis of total N-terminal pro-B type natriuretic peptide (total NT-proBNP) and of a panel of biomarkers; (2) to assess iwhether the extent of glycosylation affects the relation of NT-proBNP with AF.

**Methods:** In a sub-study of the GISSI-AF trial on 382 patients in sinus rhythm with a history of AF, echocardiographic variables and eight circulating biomarkers were serially assayed over one year. The relations between circulating baseline biomarkers and AF and the risk of CV events, were assessed by Cox-analysis models adjusting the first by clinical variables, the second by clinical variables and the echocardiographic left-atrial-minimum-volume-index (LAVI_min_).

**Results:** Over a median follow-up of 365 days, 203/382 patients (53.1%) had at least one recurrence of AF and 16.3% were hospitalized for cardiovascular (CV) reasons. Total NT-proBNP, NT-proBNP, angiopoietin 2 (Ang2), myosin binding protein (MyBPC3) and bone morphogenic protein-10 (BMP-10) were strongly associated to ongoing AF. Natriuretic peptides and MyBPC3 predicted recurrent AF but this lost significance after adjustment for LAVI_min_. NT-proBNP and Ang2 predicted CV hospitalization even after adjustment for LAVI_min_, HR95%CI: 2.20 [1.02-4.80] and 5.26 [1.16-23.79].

**Conclusions:** The association of AF recurrence with the novel biomarker total NT-proBNP, is similar to that of NT-proBNP, suggesting no influence of glycosylation. Ang2, MyBPC3 and BMP10 were strongly associated with AF, indicating a possible role of extracellular matrix and myocardial injury. Abstract-words=233

**Key messages:** *What is already known on this subject?:* *It is still complicated to predict the recurrence of AF in patients in sinus rhythm with a recent history of AF*. Though several biomarkers have been associated with AF, few of them have proved to be independent predictors for recurrent AF or cardiovascular (CV) events. Their predictive sensitivity and specificity is modest at best. Previous studies showed that NT-proBNP was possibly the strongest predictor of recurrent AF and CV hospitalization. Natriuretic peptides circulate to a large extent as glycosylated molecules and a novel assay is now available to measure the glycosylated and non-glycosylated NT-proBNP in plasma, the total NT-proBNP. The extent of glycosylation varies in different diseases.

*What might this study add?:* No studies have assessed (a) the extent of NT-proBNP glycosylation in AF, or (b) the association and predictive value in patients with AF of total NT-proBNP. A multimarker approach, ratter than one based on a single biomarker, might predict AF better. The relation with AF of the novel biomarker, total NT-proBNP, is as strong as that of NT-proBNP, suggesting no-influence of glycosylation. Two biomarkers, MyBPC3, secreted few minutes after myocardial injury and Ang-2, involved in inflammation and coagulation, were strongly associated to AF.

*How might this impact on clinical practice?:* The identification of novel circulating biomarkers could have a direct impact on clinical practice when predicting the occurrence of AF, but unfortunately current data do not allow predictions based on biomarkers. The associations of different biomarkers with ongoing AF may cast light on the mechanisms of triggering and maintenance of AF.

*Strengths and limitations of this study:* The data came from to a multicenter randomized clinical trial with available concomitant serial echocardiographic and circulating biomarkers recorded and evaluated centrally, hence with minimal bias; AF recurrence during a 12-month follow up was checked weekly by trans-telephonic electrocardiographic monitoring, and with 12-lead ECG every six months. A comparative analysis of total NT-proBNP with other novel biomarkers and echocardiographic variables has never been done so far. The possible added value of total NT-proBNP to the benchmark biomarker NT-proBNP was assessed on the basis of different dimensions of performance, as recently proposed for new biomarkers. The main limitations are (1) the relatively small numbers of patients with AF during follow-up visits, (2) the very low prevalence of patients with other cardiac diseases such as coronary artery disease and heart failure, and (3) consequently, the low incidence of clinical events in one-year follow-up.

## INTRODUCTION

Knowledge about atrial fibrillation (AF) has been steadily increasing over the last two decades together with awareness that this arrhythmia represents an important health problem [1]. In particular is a still unresolved issue, the identification of clinical and biohumoral predictive markers for AF recurrence in patients in sinus rhythm but with a recent history of AF [2–4]. Within the GISSI-AF bio-humoral sub-study the plasma concentrations of nine different biomarkers at baseline and over one-year follow-up were measured in 382 patients (5). In a multivariable model also including also echocardiographic variables, NT-proBNP was the strongest predictor of recurrent AF, duration of recurrence and cardiovascular (CV) hospitalization [5]. N-terminal pro-B type natriuretic peptide (NT-proBNP) and BNP are produced in equimolar amounts in the cardiomyocytes in response to increased wall stretch, volume overload and ischemia (6-8); BNP but not NT-proBNP has physiological activity.

There are nine known O-glycosylation sites on proBNP and NT-proBNP [9]. Up to 80% of circulating NT-proBNP in HF patients is glycosylated in the central region of the molecule. Commercial NT-proBNP ELISA contains antibodies directed to epitopes located in the central region of NT-proBNP so it underestimates the circulating concentrations of NT-proBNP when these sites are glycosylated [10,11].

An assay for “total NT-proBNP”, which also “sees” circulating glycosylated species, has recently been developed [5,12,13]. Patients eligible for the GISSI-AF trial were at high risk of recurrent AF, but had to be in sinus rhythm at inclusion [14,15]. Circulating biomarkers weakly predict AF recurrence, the most promising being NT-proBNP and high-sensitivity troponin T (hs-cTnT) [12]. The subgroup of patients in the echocardiographic and bio-humoral sub-study of GISSI-AF was deemed adequate to test independently two features of circulating biomarkers in AF, with special focus on the novel candidate biomarker, total NT-proBNP: 1) the association of a biomarker with AF, when a blood sample is taken when AF is present, and 2) the predictive power of a biomarker, meaning the relation between its concentration when the patient is in sinus rhythm and the probability of new AF or incident CV adverse events.

## METHODS

The GISSI-AF trial was a double-blind randomized placebo-controlled multicenter trial that enrolled 1442 patients in sinus rhythm with a history of AF (two or more episodes of symptomatic ECG-documented AF in the previous six months) or successful cardioversion, electrical or pharmacologic between 14 days and 48 hours before randomization). The rationale, design, and results of the trial have already been published (*Clinical Trials*.*gov identifier: NCT00376272*) [14,15].

A total of 382 patients (26.5%) from 36 centers participated in a sub-study with serial echocardiographic and biohumoral examining at baseline and at 12-month follow-up (end of study). Core laboratories read all echocardiograms and analyzed all blood samples under blind conditions. Biomarkers were assayed in a laboratory at Roche Diagnostics, Penzberg, Germany.

### Echocardiography, assays of circulating biomarkers and detection of AF recurrence

Details on echocardiography, the biomarkers characteristics and assays and methods for detection of recurrent AF are given in *Online supplementary appendix 1*.

### Statistical methods

Continuous variables are expressed as mean±SD if normally distributed or median and interquartile range [IQR], as appropriate; categorical variables were reported as absolute numbers and percentages. Differences between groups were analysed with one-way ANOVA, Kruskal-Wallis test or χ2 test, as appropriate. The correlations between circulating biomarkers and echocardiographic variables were calculated with *Spearman’s* rank correlation coefficient. The discriminatory ability of each biomarker for recurrent AF was assessed by ROC analysis. Restricted cubic splines were employed to assess the continuous relationship between biomarkers and the log relative hazard for AF recurrence. Kaplan Meier, then log-rank analysis was used to evaluate differences in AF-free survival.

Three different Cox proportional hazards models were employed to assess the association between total NT-proBNP and predictive value for incident AF and for clinical events: *Model 1*, adjusted for sex and more than two recurring AF episodes within six months from inclusion; *Model 2*, where LAVI_min_ was added to *Model 1*; and *Model 3* where total NT-proBNP or NT-proBNP were was added to Model 2. LAVI_min_ was chosen among the echocardiogaphic variables for its predictive power for incident AF [16], according to a parsimonious approach. Biomarkers and LAVI_min_ were entered as continuous variables (1 point increase for biomarkers and LAVI_min_).

All probability values are two-tailed and p <0.05 was considered significant.

## RESULTS

### Patients

The biomarker sub-study included 382 patients and 1070 plasma samples were analyzed. The baseline characteristics of this population were similar to those of the 1442 patients enrolled in the main GISSI-AF study [15]. In brief, age was 67.6 ± 9.1 years, 142 (37.1%) were women, 154 (0.8%) presented ≥2 episodes of AF in the previous six months and 336 (88.0%) patients had undergone cardioversion for AF in the previous two weeks. A history of hypertension was present in 334 (84.8%) and heart failure (HF) or LVEF<40% in 42 (11.0%) patients. (*Online supplementary appendix 2*). The study treatment, valsartan was given to 184 patients (48.7%), ACE-inhibitors to 206 (53.9%), beta-blockers to 114 (29.8%) and aldosterone blockers to 20 (5.2%).

During follow-up (median 365 days, range 5-373 days), 203 (53.1%) patients had at least one newly diagnosed episode of AF and 113 (29.6%) had more than one. The median number of episodes of recurrent AF per patient was 3 (range 2-27). During the one-year follow-up one patient died, and the incidence of hospitalizations for any reason was 18.7% and 16.3% for CV reasons.

As there were no significant differences between valsartan and placebo for any of the circulating biomarkers and echocardiographic variables, the whole cohort of 382 patients was analysed, irrespective of the treatment randomized.

### Plasma concentrations of circulating biomarkers at baseline and during follow-up

Concentrations of total NT-proBNP and of the other biomarkers at each study visit are shown in *Online supplementary appendix 3*, with their reference ranges; medians were generally within the reference range except for total NT-proBNP, NT-pro BNP, IGFBP7 and for GDF-15, where the levels were slightly above the normal range. Median concentrations of all biomarkers were either stable or decreased slightly over the 12-month follow-up.

### Relationship between circulating biomarkers and echocardiographic variables

Patients presented the following echocardiographic characteristics: LAVI_max_ 42.6±14.7 ml/m^2^, LAVI_min_ 23.3± 12.3 mL/m^2^, LAEF 47.5±13.3%, LVEF 60.5±12.3 %; 205/270 (53.7%) patients had grade 1 mitral regurgitation. The E/e’ ratio was available for 108 patients, with a mean of 12.6±8.6 (*Online supplementary appendix 2*). LAEF and LVEF were inversely correlated and LAVI_max_ and LAVI_min_ were positively correlated with all circulating biomarkers. The highest Spearman’s rank coefficients were for total NT-proBNP, followed by NT-proBNP, Ang-2, BMP10 and MyBPC3, Table 1. E/e’ appeared to be unrelated to any of the circulating biomarkers.

**Table 1.** Correlation between echocardiographic left atrial and ventricular variables and circulating biomarkers.

|  |  | LAVI<sub>max</sub> | LAVI<sub>min</sub> | LAEF | E/e' | LVEF |
| --- | --- | --- | --- | --- | --- | --- |
| Total NT-proBNP (pg/mL) | baseline | 0.51 | 0.56 | -0.50 | -0.17 | 0.24 |
|  | 6 months | 0.34 | 0.40 | -0.42 | -0.14 | 0.35 |
|  | 12 months | 0.40 | 0.49 | -0.42 | -0.21 | 0.26 |
| NT-proBNP (pg/mL) | baseline | 0.48 | 0.54 | -0.48 | -0.21 | 0.28 |
|  | 6 months | 0.28 | 0.37 | -0.39 | -0.21 | 0.32 |
|  | 12 months | 0.39 | 0.46 | -0.39 | -0.25 | 0.26 |
| Ang2 (ng/mL) | baseline | 0.36 | 0.41 | -0.37 | -0.15 | 0.15 |
|  | 6 months | 0.16 | 0.21 | -0.25 | -0.12 | 0.27 |
|  | 12 months | 0.21 | 0.28 | -0.23 | -0.20 | 0.09 |
| BMP10 (ng/ml) | baseline | 0.29 | 0.32 | -0.26 | 0.07 | 0.18 |
|  | 6 months | 0.19 | 0.20 | -0.20 | 0.07 | 0.24 |
|  | 12 months | 0.19 | 0.21 | -0.19 | 0.06 | 0.22 |
| DKK3 (ng/mL) | baseline | 0.30 | 0.29 | -0.23 | -0.04 | 0.11 |
|  | 6 months | 0.22 | 0.24 | -0.24 | -0.08 | 0.18 |
|  | 12 months | 0.22 | 0.25 | -0.17 | -0.05 | 0.19 |
| ESM1 (ng/mL) | baseline | 0.27 | 0.27 | -0.22 | 0.01 | 0.21 |
|  | 6 months | 0.17 | 0.13 | -0.10 | 0.03 | 0.18 |
|  | 12 months | 0.24 | 0.22 | -0.11 | -0.07 | 0.20 |
| FABP3 (ng/mL) | baseline | 0.08 | 0.16 | -0.23 | 0.07 | 0.06 |
|  | 6 months | 0.01 | 0.09 | -0.21 | 0.05 | 0.03 |
|  | 12 months | 0.07 | 0.17 | -0.21 | 0.07 | 0.06 |
| FGF23 (ng/ml) | baseline | 0.16 | 0.21 | -0.19 | -0.06 | 0.18 |
|  | 6 months | 0.05 | 0.15 | -0.22 | -0.03 | 0.28 |
|  | 12 months | 0.07 | 0.16 | -0.21 | -0.05 | 0.34 |
| GDF15 (pg/mL) | baseline | 0.23 | 0.32 | -0.34 | -0.05 | 0.28 |
|  | 6 months | 0.19 | 0.21 | -0.23 | -0.01 | 0.23 |
|  | 12 months | 0.22 | 0.26 | -0.28 | -0.06 | 0.31 |
| IGFBP7 (ng/mL) | baseline | 0.29 | 0.31 | -0.28 | -0.03 | 0.16 |
|  | 6 months | 0.15 | 0.21 | -0.23 | -0.02 | 0.27 |
|  | 12 months | 0.20 | 0.25 | -0.20 | -0.01 | 0.23 |
| MYBPC3 (pg/ml) | baseline | 0.41 | 0.44 | -0.40 | -0.23 | 0.40 |
|  | 6 months | 0.33 | 0.40 | -0.44 | -0.15 | 0.45 |
|  | 12 months | 0.36 | 0.43 | -0.40 | -0.20 | 0.46 |
| LAVI<sub>max</sub> |  | 1.00 | 0.85 | -0.44 | -0.26 | 0.14 |
| LAVI<sub>min</sub> |  | 0.85 | 1.00 | -0.77 | -0.28 | 0.16 |
| LAEF |  | -0.44 | -0.77 | 1.00 | 0.28 | -0.20 |
| E/e' |  | -0.26 | -0.28 | 0.28 | 1.00 | -0.04 |
| LVEF |  | 0.14 | 0.16 | -0.20 | -0.04 | 1.00 |

### Relation between circulating biomarkers and ongoing AF

During the one-year follow-up, 34 and 45 patients presented AF, respectively at 6- and 12-months. Concentrations of total NT-proBNP, NT-proBNP, Ang2, BMP10, DKK3, FGF-23 and MyBPC3 were significantly higher in those with AF at each study visit. Natriuretic peptides and Ang2 had by far the largest significant increase in the presence of AF (ranging from 6.5×10^−15^ to 1.7×10^−5^), **Table 2**.

**Table 2.**
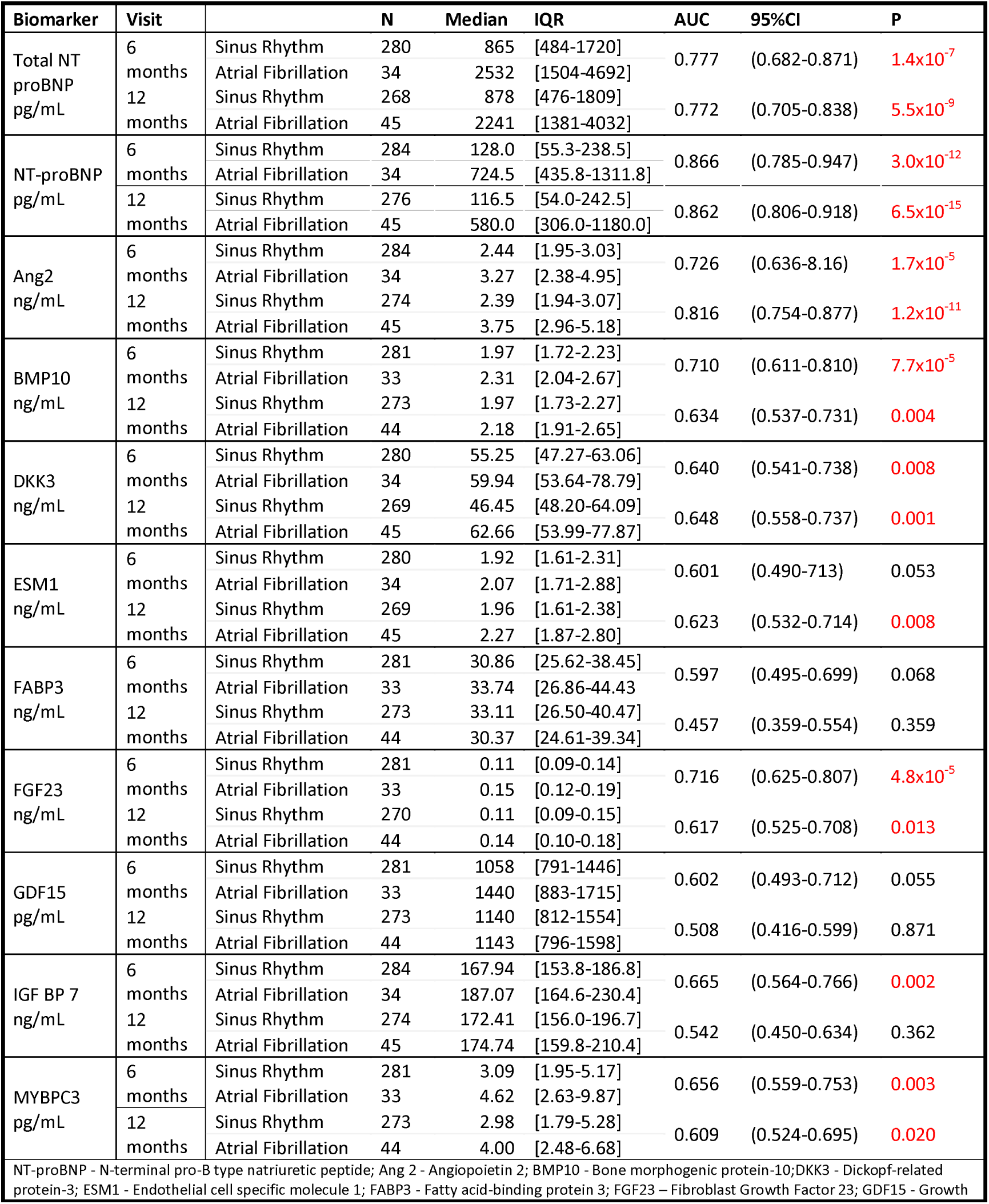
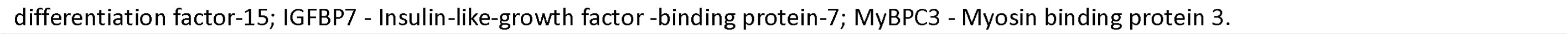
Concentrations of circulating biomarkers at baseline and in patients with atrial fibrillation/sinus rhythm recorded by ECG during scheduled visits at 6 and 12 months.

To assess the strength of the relation between biomarkers and ongoing AF (i.e. biomarkers assayed while an AF rhythm was recorded), ROC analyses were done. The AUCs were highest at 6 and 12 months for total NT-proBNP (AUC_6M_ 0.78 and AUC_12M_ 0.77), NT-proBNP (AUC_6M_ 0.87 and AUC_12M_ 0.86), Ang2 (AUC_6M_ 0.73 and AUC_12M_ 0.82), and BMP10 (AUC_6M_ 0.71 and AUC_12M_ 0.63), **Table 2**).

For the ratio of total NT-proBNP/NT-proBNP AUCs improved better than for each natriuretic peptide alone (AUC_6M_ 0.93 and AUC_12M_ 0.91), suggesting lowers levels of glycosylated NT-proBNP in ongoing AF. Baseline biomarkers values in univariate analysis were significantly associated with ongoing AF, and this association remained significant after adjustment for clinical variables (*Model 1*). When echocardiographic variables (*Model 2*) and total NT-proBNP or NT proBNP (*Model 3*), were added to the model the statistical significance of the associations was markedly reduced in the first case, **Table 3** and disappeared in the second (data not shown).

**Table 3.**
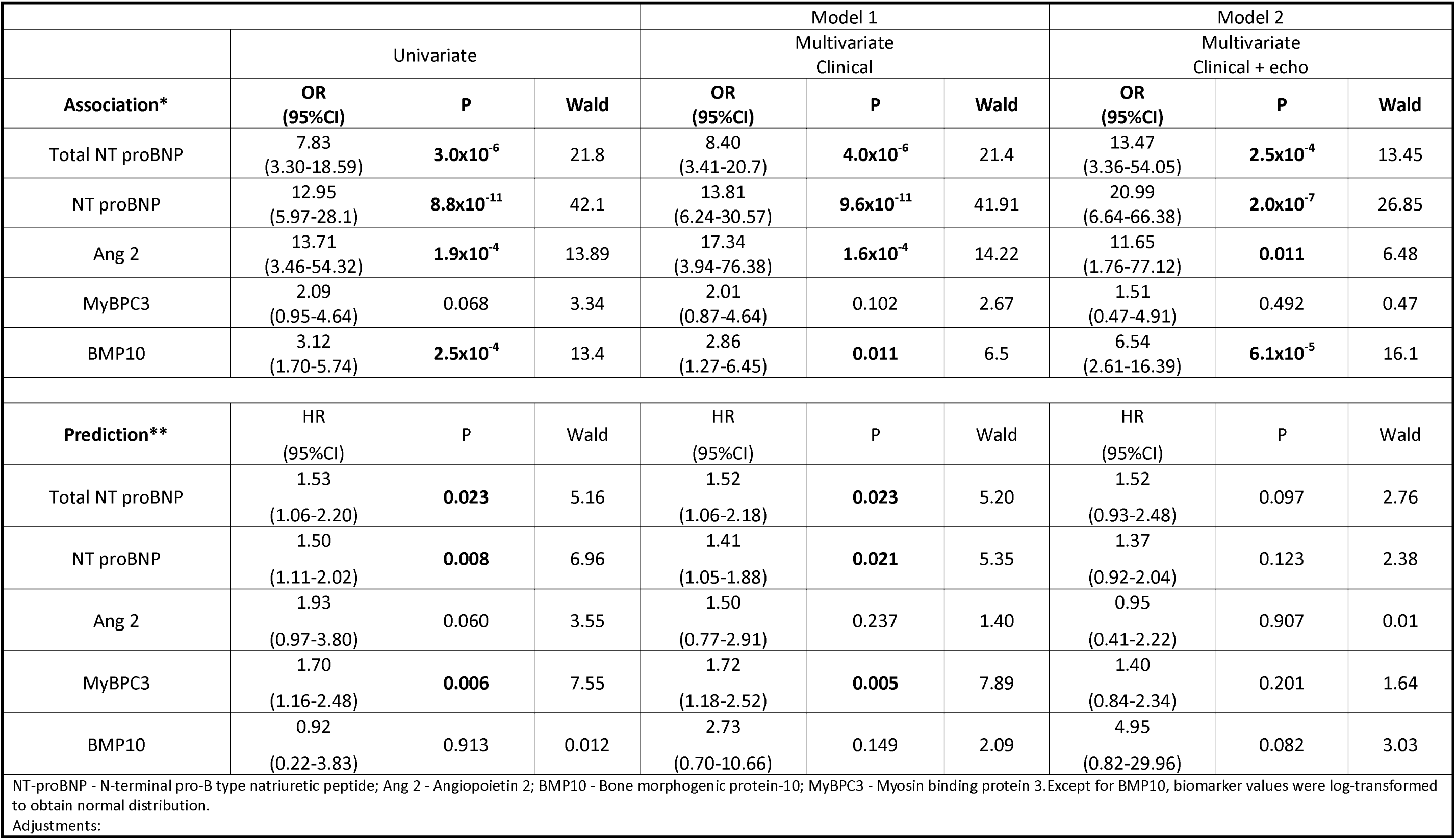
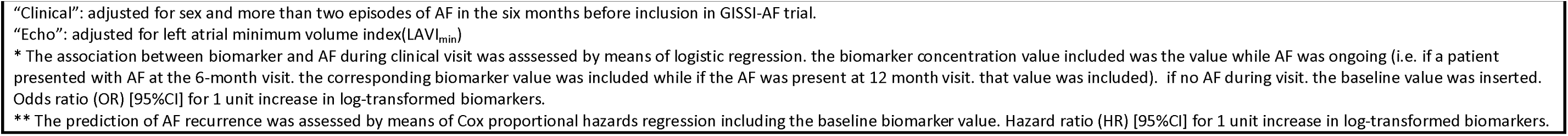
Multivariate regression models for biomarkers and atrial fibrillation prediction and association.

|  |  |  |  | Model 1 |  |  | Model 2 |  |  |
| --- | --- | --- | --- | --- | --- | --- | --- | --- | --- |
|  | Univariate |  |  | Multivariate Clinical |  |  | Multivariate Clinical + echo |  |  |
| Association* | OR<br>(95%CI) | P | Wald | OR<br>(95%CI) | P | Wald | OR<br>(95%CI) | P | Wald |
| Total NT proBNP | 7.83<br>(3.30-18.59) | **3.0x10<sup>-6</sup>** | 21.8 | 8.40<br>(3.41-20.7) | **4.0x10<sup>-6</sup>** | 21.4 | 13.47<br>(3.36-54.05) | **2.5x10<sup>-4</sup>** | 13.45 |
| NT proBNP | 12.95<br>(5.97-28.1) | **8.8x10<sup>-11</sup>** | 42.1 | 13.81<br>(6.24-30.57) | **9.6x10<sup>-11</sup>** | 41.91 | 20.99<br>(6.64-66.38) | **2.0x10<sup>-7</sup>** | 26.85 |
| Ang 2 | 13.71<br>(3.46-54.32) | **1.9x10<sup>-4</sup>** | 13.89 | 17.34<br>(3.94-76.38) | **1.6x10<sup>-4</sup>** | 14.22 | 11.65<br>(1.76-77.12) | **0.011** | 6.48 |
| MyBPC3 | 2.09<br>(0.95-4.64) | 0.068 | 3.34 | 2.01<br>(0.87-4.64) | 0.102 | 2.67 | 1.51<br>(0.47-4.91) | 0.492 | 0.47 |
| BMP10 | 3.12<br>(1.70-5.74) | **2.5x10<sup>-4</sup>** | 13.4 | 2.86<br>(1.27-6.45) | **0.011** | 6.5 | 6.54<br>(2.61-16.39) | **6.1x10<sup>-5</sup>** | 16.1 |
| Prediction** | HR<br>(95%CI) | P | Wald | HR<br>(95%CI) | P | Wald | HR<br>(95%CI) | P | Wald |
| Total NT proBNP | 1.53<br>(1.06-2.20) | **0.023** | 5.16 | 1.52<br>(1.06-2.18) | **0.023** | 5.20 | 1.52<br>(0.93-2.48) | 0.097 | 2.76 |
| NT proBNP | 1.50<br>(1.11-2.02) | **0.008** | 6.96 | 1.41<br>(1.05-1.88) | **0.021** | 5.35 | 1.37<br>(0.92-2.04) | 0.123 | 2.38 |
| Ang 2 | 1.93<br>(0.97-3.80) | 0.060 | 3.55 | 1.50<br>(0.77-2.91) | 0.237 | 1.40 | 0.95<br>(0.41-2.22) | 0.907 | 0.01 |
| MyBPC3 | 1.70<br>(1.16-2.48) | **0.006** | 7.55 | 1.72<br>(1.18-2.52) | **0.005** | 7.89 | 1.40<br>(0.84-2.34) | 0.201 | 1.64 |
| BMP10 | 0.92<br>(0.22-3.83) | 0.913 | 0.012 | 2.73<br>(0.70-10.66) | 0.149 | 2.09 | 4.95<br>(0.82-29.96) | 0.082 | 3.03 |
| NT-proBNP - N-terminal pro-B type natriuretic peptide; Ang 2 - Angiotensin 2; BMP10 - Bone morphogenetic protein-10; MyBPC3 - Myosin binding protein 3. Except for BMP10, biomarker values were log-transformed to obtain normal distribution.<br>Adjustments: |  |  |  |  |  |  |  |  |  |

“Clinical”: adjusted for sex and more than two episodes of AF in the six months before inclusion in GISSI-AF trial.
“Echo”: adjusted for left atrial minimum volume index(LAVI<sub>min</sub>)
\* The association between biomarker and AF during clinical visit was assessed by means of logistic regression. the biomarker concentration value included was the value while AF was ongoing (i.e. if a patient presented with AF at the 6-month visit. the corresponding biomarker value was included while if the AF was present at 12 month visit. that value was included). if no AF during visit. the baseline value was inserted. Odds ratio (OR) [95%CI] for 1 unit increase in log-transformed biomarkers.
\*\* The prediction of AF recurrence was assessed by means of Cox proportional hazards regression including the baseline biomarker value. Hazard ratio (HR) [95%CI] for 1 unit increase in log-transformed biomarkers.

On the basis of the association with AF rhythm during follow-up, assessed by multiple tests (see **Tables 2 and 3**), the following biomarkers were selected to evaluate their predictive abilities for AF recurrence: Ang2, BMP10, MyBPC3 and Total NT-proBNP as well as the benchmark marker NT-proBNP and the total NT-proBNP/NT-proBNP ratio.

### Circulating biomarkers as predictors of AF recurrence and clinical outcomes

During the one-year follow-up, 203 (53.1%) patients experienced at least one episode of AF recurrence, mostly identified in telemetric device recordings. Clinical characteristics and treatments of patients with at least one AF recurrence and with any recurrence were previously published [12] (see also *Online supplementary appendix 3*). Males and patients with a history of more than two episodes of AF in the six months prior to inclusion in the study were more likely to suffer at least one AF recurrence. Baseline concentrations of biomarkers in patients with or without AF recurrence were similar: only MyBPC3 showed a borderline-significance (median [IQR], 3.69 pg/mL [2.29-6.35], vs 3.19 pg/mL [1.97-6.13], p=0.052), **Table 4**.

**Table 4.**
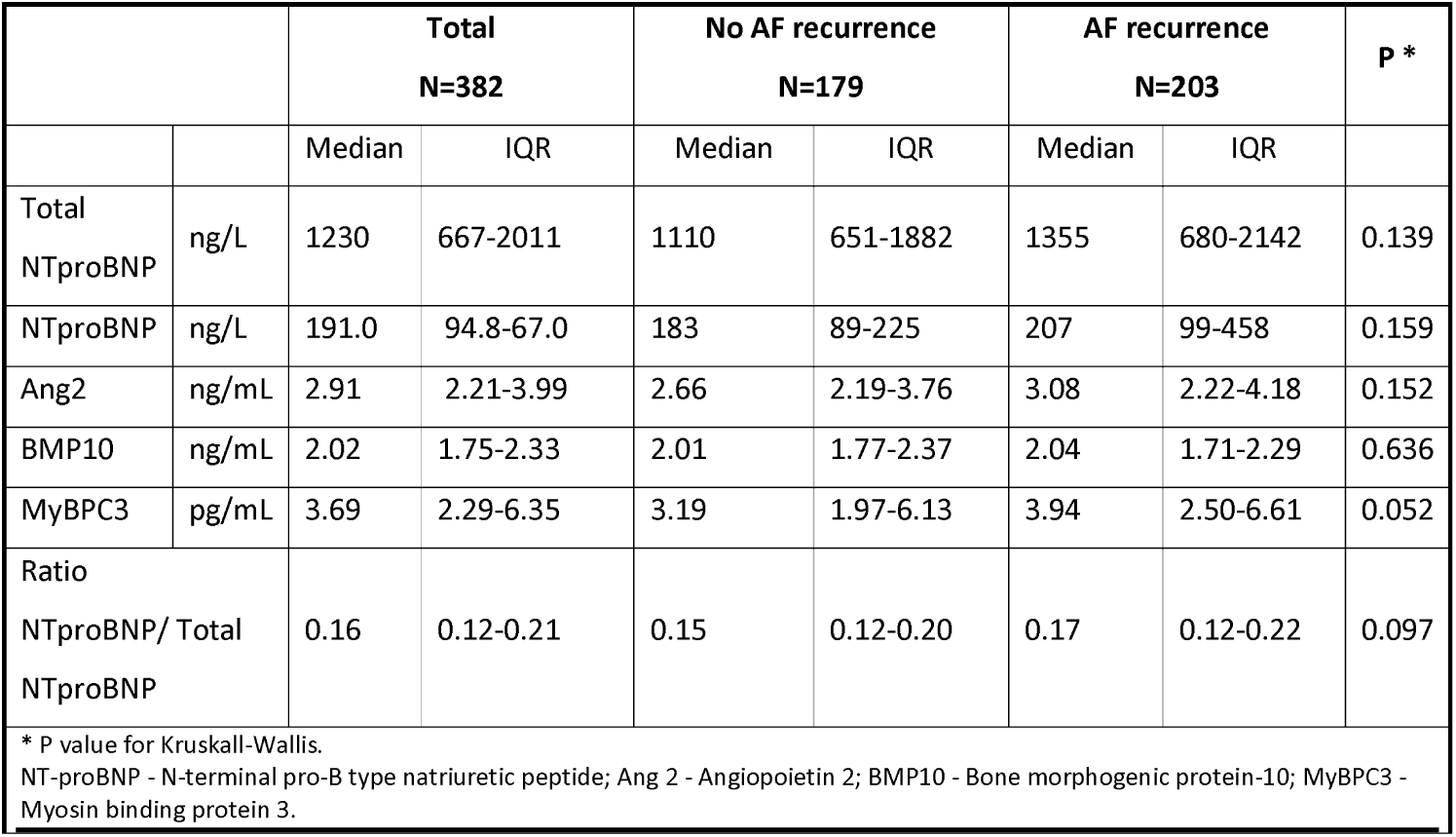
Concentrations of the circulating biomarkers at baseline in patients with and without AF recurrence.

Kaplan Meier curves for AF recurrence in relation to baseline biomarker concentration below and above the median indicated that curves started to diverge within the first week for total NT-proBNP, NT-proBNP and at one month for Ang2 and MyBPC3, **Figure 1**. Significant differences from the median were reached only by total NT-proBNP and MyBPC3.

**Figure 1.**
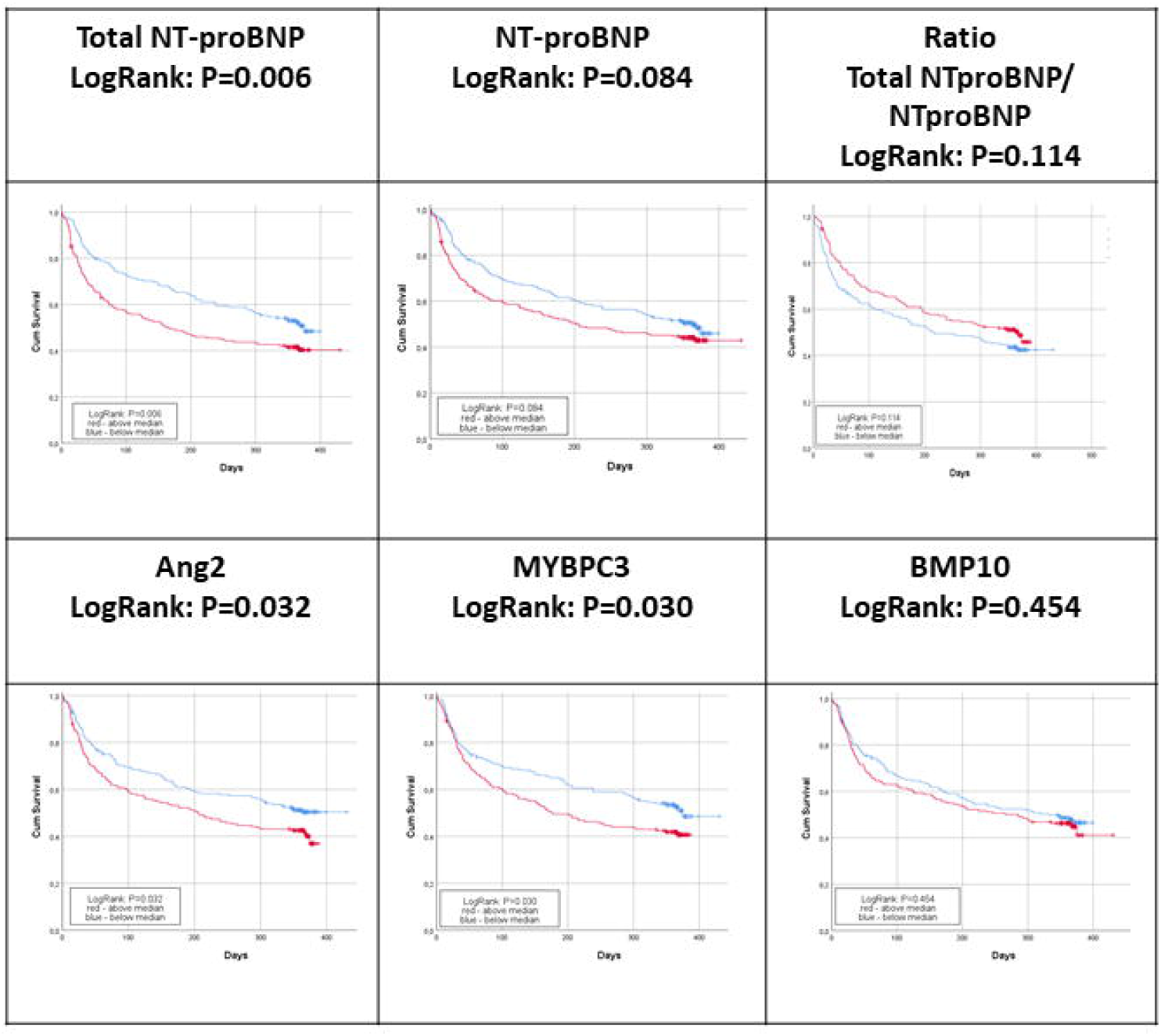
Restricted cubic spline depicting the continuous biomarker concentrations in relation to the risk of time to first AF recurrence. Total NT-proBNP: 2=5.91, p=0.052; NT-proBNP: 2=12.05, p=0.002, Total NT-proBNP/NT-proBNP: 2=5.69, p=0.058; MyBPC3: 2=7.01, p=0.030; Ang2: 2=4.46, p=0.107 and BMP1: 2=0.83, p=0.363. The continuous line indicates the central log relative hazard and the shaded area the 95% confidence intervals. Biomarkers were transformed to a natural logarithmic scale for this study endpoint. P value for factor chi-square from Wald statistics.

**Figure 2** shows the relationship, as restricted cubic splines, between AF recurrence and baseline concentrations of each biomarker. Total NT-pro BNP and total NT-proBNP/NT-proBNP were weakly associated with incident AF (p=0.052 and 0.058), but NT-proBNP and MyBPC3 reached statistical significance (p= 0.002 and 0.030 respectively).

ROC for recurrent AF yielded AUCs ranging from 0.54 to 0.59 for the different molecules, not reaching statistical significance (data not shown).

**Figure 2.**
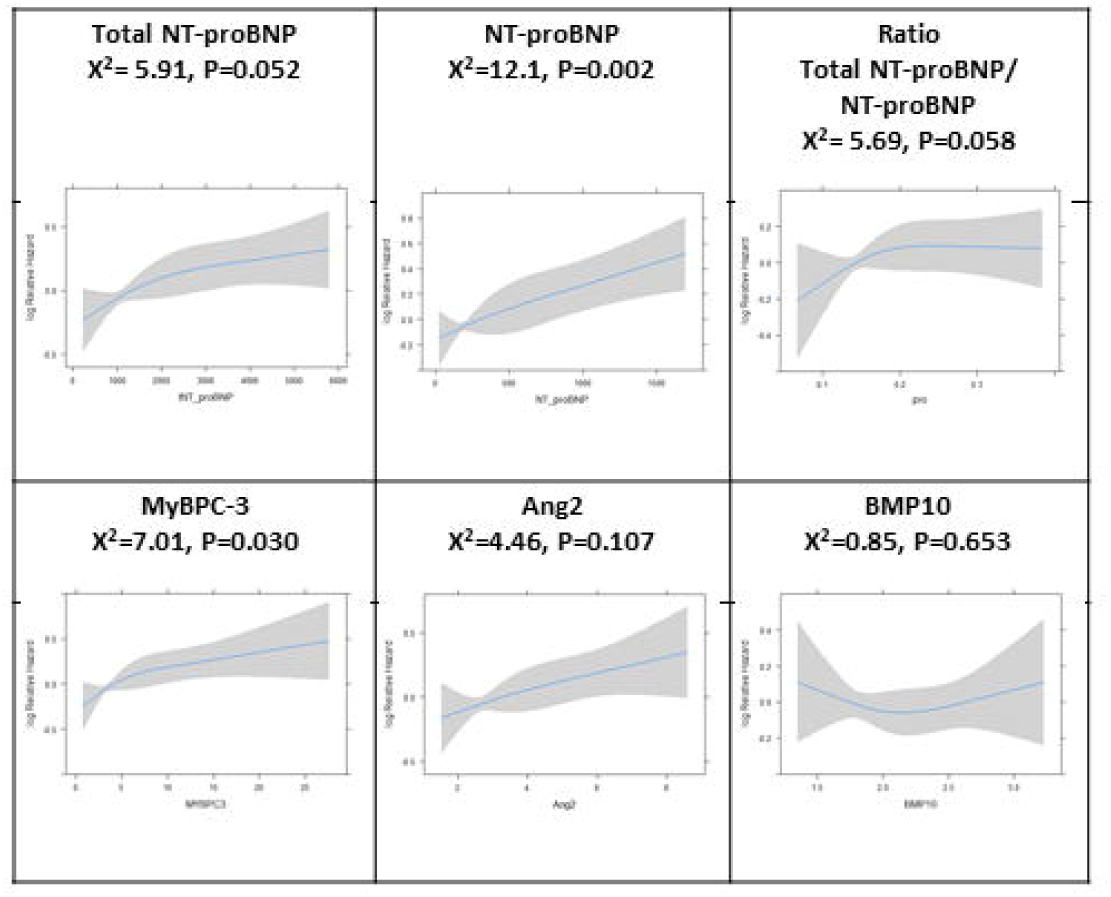
Kaplan-Meier curves of first recurrence of AF by median baseline biomarker concentration. Log rank test for: total NT-proBNP, p=0.006; NT-proBNP, p=0.084; total NT-proBNP/ NT-proBNP, p=0.114; MyBPC3, p=0.030; Ang2, p=0.032 and BMP10, p=0.454.

The predictive power for recurrent AF of baseline concentrations of total NT-proBNP, NT-proBNP, Ang2, MyBPC3 and BMP10 was also assessed with Cox proportional hazard models. In univariate analysis all four biomarkers significantly predicted AF recurrence. However, after adjustment for clinical variables (i.e. male sex and more than two episodes of AF within six months before inclusion in the GISSI-AF study, *Model 1*) analyzed biomarkers were still significantly associated to AF recurrence except for Ang2, **Table 3**. When echocardiographic variables were added to the model (*Model 2*) only Ang2 was weakly significant, while all significance disappeared after inclusion of total NT-proBNP or NT-proBNP (*Model 3*) (data not shown).

### Hospitalization for CV reasons

Total NT-proBNP was associated with hospitalizations for CV reasons both in univariate [HR [95%CI]: 3.02 [1.45-6.28], p=0.003] and multivariate Model 1. Only NT-proBNP and Ang2 were still significantly associated with CV hospitalizations after inclusion of LAVI_min_, **Table 5**. Statistical significance disappeared after inclusion of total NT-proBNP or NT-proBNP (*Model 3*) (data not shown).

**Table 5.**
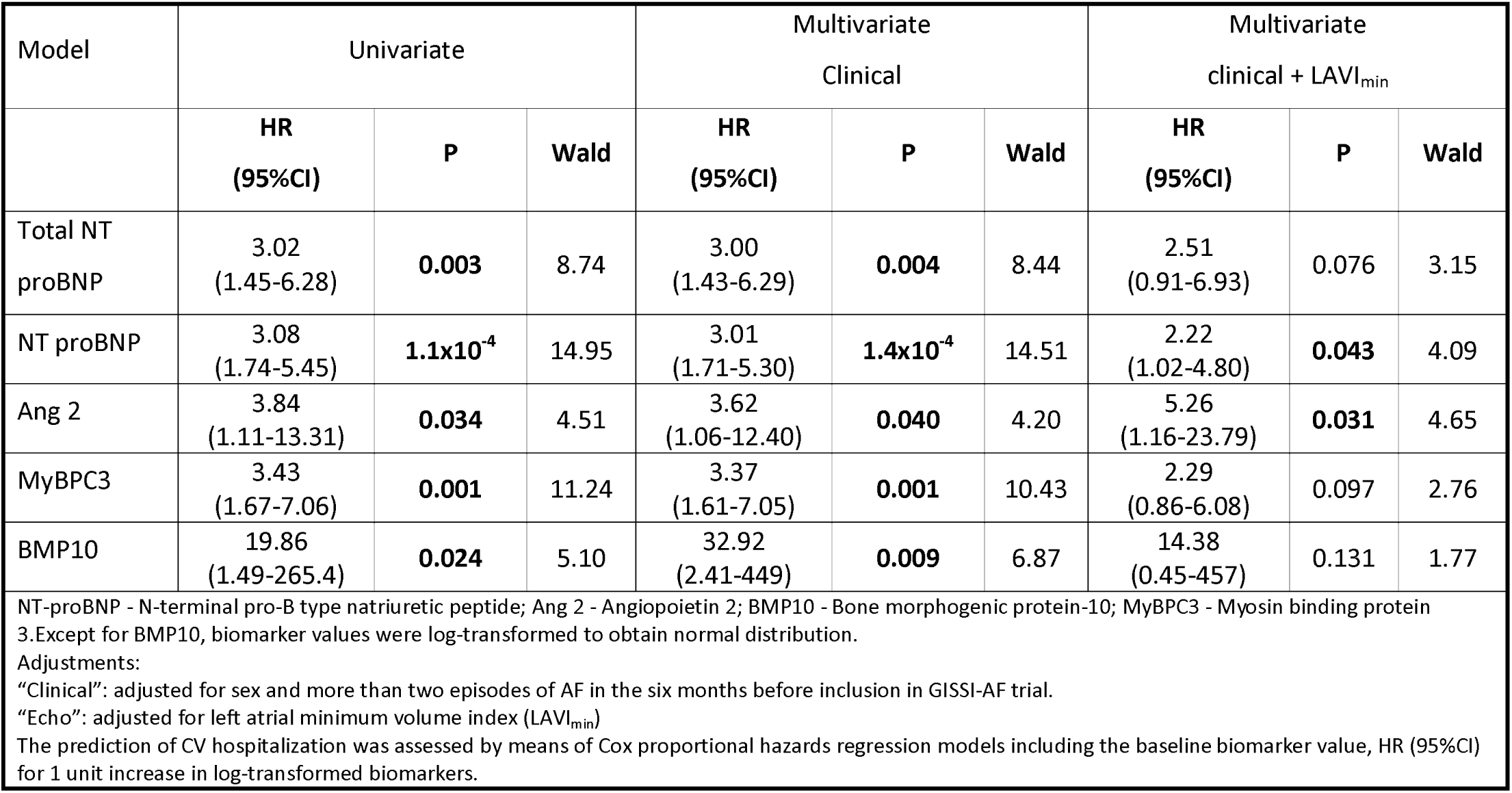
Multivariate Cox proportional hazards regression models for biomarkers and hospitalizations for CV reasons.

| Model | Univariate |  |  | Multivariate<br>Clinical |  |  | Multivariate<br>clinical + LAVI<sub>min</sub> |  |  |
| --- | --- | --- | --- | --- | --- | --- | --- | --- | --- |
|  | HR<br>(95%CI) | P | Wald | HR<br>(95%CI) | P | Wald | HR<br>(95%CI) | P | Wald |
| Total NT<br>proBNP | 3.02<br>(1.45-6.28) | **0.003** | 8.74 | 3.00<br>(1.43-6.29) | **0.004** | 8.44 | 2.51<br>(0.91-6.93) | 0.076 | 3.15 |
| NT proBNP | 3.08<br>(1.74-5.45) | **1.1x10<sup>-4</sup>** | 14.95 | 3.01<br>(1.71-5.30) | **1.4x10<sup>-4</sup>** | 14.51 | 2.22<br>(1.02-4.80) | **0.043** | 4.09 |
| Ang 2 | 3.84<br>(1.11-13.31) | **0.034** | 4.51 | 3.62<br>(1.06-12.40) | **0.040** | 4.20 | 5.26<br>(1.16-23.79) | **0.031** | 4.65 |
| MyBPC3 | 3.43<br>(1.67-7.06) | **0.001** | 11.24 | 3.37<br>(1.61-7.05) | **0.001** | 10.43 | 2.29<br>(0.86-6.08) | 0.097 | 2.76 |
| BMP10 | 19.86<br>(1.49-265.4) | **0.024** | 5.10 | 32.92<br>(2.41-449) | **0.009** | 6.87 | 14.38<br>(0.45-457) | 0.131 | 1.77 |
| NT-proBNP - N-terminal pro-B type natriuretic peptide; Ang 2 - Angiopoietin 2; BMP10 - Bone morphogenic protein-10; MyBPC3 - Myosin binding protein<br>3.Except for BMP10, biomarker values were log-transformed to obtain normal distribution.<br>Adjustments:<br>"Clinical": adjusted for sex and more than two episodes of AF in the six months before inclusion in GISSI-AF trial.<br>"Echo": adjusted for left atrial minimum volume index (LAVI<sub>min</sub>)<br>The prediction of CV hospitalization was assessed by means of Cox proportional hazards regression models including the baseline biomarker value, HR (95%CI)<br>for 1 unit increase in log-transformed biomarkers. |  |  |  |  |  |  |  |  |  |

## DISCUSSION

While the association of a biomarker with AF may provide useful mechanistic insights on the disease, a biomarker that can tell the doctor in advance what is the risk of an individual patient having new episodes of AF is clinically relevant. That is why we comparatively assessed, for the first time, the strength of the association and the predictive power for AF of the plasma concentrations of total NT-proBNP in a cohort of patients in sinus rhythm with a history of AF, over one year of follow-up at high risk of recurrence.

Median Total NT-proBNP in plasma were 6.6, 7.2 and 6.8 times higher than NT-proBNP in patients with AF rhythm at baseline at six and 12 months follow-up, Table 1. This indicates that in these patients BNP is extensively glycosylated. In patients with acute dyspnea, Røsjø et al. [17] reported nearly two-fold higher levels of total NT-proBNP than NT-proBNP in HF and non-HF patients. Although both natriuretic peptides were predictive of death in HF patients at a median of 814 days follow-up, the de-glycosylated total NT proBNP concentration was stronger in predicting death (Wald, Chi^2^ 24.1) than NT proBNP (Wald, Chi^2^ 14.3). In the present study, there were 42 patients with clinically diagnosed HF or LVEF<40% (11%); this small number of patients does not allow an analysis of AF stratified by HF, although the incidence of AF was no different in patients with HF (12.3%) from patients without (9.5%, p=0.38). Nonetheless, circulating biomarkers were significantly higher in patients with HF, independently of AF. The only exception was BMP10, apparently independent of HF (*Online supplementary appendix 4*).

Although it was not the main aim of this study, the association and predictive value of MyBPC3 for AF in patients at high risk of recurrence is a new finding. However, its significance in adjusted multivariable models is borderline. MyBPC3 showed a strong correlation with LAVI_min_, close to natriuretic peptides (**Table 2**). MyBPC3 is a new biomarker of myocardial necrosis reported to have an overall performance comparable to troponin assays in the diagnosis of acute myocardial infarction [18]. Higher baseline hs-cTnT in previous published results of the GISSI-AF biomarker substudy [12] were independently associated with AF recurrence {adjusted hazard ratio (HR) [95% confidence intervals (CI) for 1 SD increment] (1.15 [1.04-1.28], P = 0.007} and to CV hospitalizations (1.19 [1.01-1.39], P = 0.03), similar to the predictive performance of MyBPC3 after adjustment for clinical variables (HR and [95% CI]) for 1 SD increment] 1.72 [1.18-2.52], P = 0.005) and 3.37 [1.61-7.05], P = 0.001). Further studies may help verify the potential of MyBPC3 as a novel predictive circulating biomarker of incident AF and related CV hospitalization. Ang2 showed a strong association with AF, similar to that of NT-proBNP, confirming the well-known pathogenic role of inflammatory activation in AF [19]. However, Ang2 had no predictive power for recurrent AF, suggesting that different players may be involved in determining the risk of new occurrence of AF. In the GISSI-AF substudy patients, similar results were reported for two other inflammatory markers, IL6 and hsCRP [13]. Like for Ang2 these two biomarkers were not associated with AF recurrence. In relation to AF, another marker closely involved in inflammation such as GDF15 is highly predictive of severity and risk in HF, but performed poorly in the present study. Nonetheless, GDF15 was reported in the ARISTOTLE large-scale trial to be a risk factor for major bleeding, mortality, and stroke in patients with permanent AF [20].

BMP10 has been very recently shown to predict recurrence of AF independently after catheter ablation in 359 patients, while NT-proBNP did not [21]. However, patients were more severely ill (47% had HF, history of stroke in 12%, NT-proBNP approximately double higher that in GISSI-AF) than those in the present study, even though absolute concentrations of BMP10 were just slightly higher.

Recently natriuretic peptides, in particular NT-proBNP levels, have been reported to be associated with inflammation [22]. This suggests a common pathophysiological mechanism for the biomarkers assessed in this study that resulted associated to AF.

Our findings of a significant association of Ang2 with the risk of CV hospitalization are in agreement with a previous study [23]. Those authors reported higher Ang2 plasma levels in patients with chronic AF than in healthy subjects, together with high levels of von Willebrand factor (vWF) and vascular endothelial growth factor (VEGF). These findings were interpreted as indicating links between endothelial damage, dysfunction and thrombogenic factors that may be risk factors for CV hospitalization. In the GISSI-AF main study the incidence of thromboembolic events was low (12/1447 patients, 0.97%) [24], so this analysis is not feasible. The ratio of total NT-proBNP/NT-proBNP did not prove better than either natriuretic peptide alone, as shown by Kaplan-Meier curves and splines (Figures 1 and 2). As previously reported, the total NT-proBNP/NT-proBNP ratio was significantly lower in patients with HF or LVEF<40% (*Online supplementary appendix 4*), suggesting less glycosylation of NT-proBNP in HF [25] with ventricular overload. This difference was not found in atrial overload [26].

### Strengths and limitations of the study

The main strengths of this study are (a) it was a multicenter randomized clinical trial with concomitant serial echocardiographic and circulating biomarkers analysed centrally; (b) trans-telephonic electrocardiographic monitoring enabled us to record and identify AF recurrence efficiently during the 12-month follow-up. Another strength is the comparative simultaneous analysis of total NT proBNP with other biomarkers that participate in different pathophysiological mechanisms, some of them assessed for the first time for AF diagnosis or as predictors of AF recurrence (Ang2, MyBPC3, BMP10). The possible added value of total NT-proBNP to the benchmark biomarker NT-proBNP was assessed from different dimensions of performance, as recently proposed for the evaluation of new biomarkers [20].

Our results do not apply to all patients with AF since at baseline the GISSI-AF population were in sinus rhythm and had a lower rate of co-morbidities than patients from other AF studies where there was mainly a high frequency of persistent or permanent AF. The low frequency of deaths after 12 months (only one) and thromboembolic events (n=3) in the GISSI-AF substudy could not be considered for the outcome analysis. The small number of patients with AF during a follow-up visit constitutes a limitation for the association of the biomarkers with the diagnosis of AF.

In conclusion, the performance of the new total NT-proBNP as a new biomarker in AF is similar to that of NT-proBNP. Other markers such as Ang2 and MyBPC3 show strong associations with ongoing AF, pointing to a pathogenic role of extracellular matrix and myocardial injury in AF.

## Data Availability

The entire database of GISSI-AF can be accessed upon reasonable request to the Steering Committee.

## Acknowledgements

We thank the patients, nurses and cardiologists who participated in the biohumoral substudy of GISSI-AF. We are grateful to Judith Baggott for language editing

## APPENDIX PARTICIPATING CENTRES AND INVESTIGATORS

*Switzerland*: Lugano (MG Rossi). *Italy*: Bagno a Ripoli (A Fazi), Bari Carbonara(O Pierfelice), Bergamo (A Gavazzi, F Taddei), Bovolone (G Rigatelli, S Boni), Bussolengo (R Trappolin), Casarano (A Muscella), Caserta (A Vetrano), Catania (M Gulizia,GM Francese), Catanzaro (F Perticone), Città di Castello (D Severini), Cremona (S Pirelli, A Spotti, M Mariani), Fidenza (P Pastori), Firenze (GM Santoro, C Minneci), NapoliFederico II (P Perrone Filardi), Palermo Cervello (L Buffa), Palermo Villa Sofia (F Ingrillì),Pavia (L Tavazzi, C Belvito), Pesaro (A Pierantozzi), Pietra Ligure (A Nicolino), Reggio Calabria (G Pulitanò, A Ruggeri, G Cutrupi), Roma (M Volpe), Saluzzo (S Reynaud), SanBonifiacio (R Rossi, E Carbonieri, E Zampieri), San Daniele del Friuli (L Mos, G Marcuzzi),San Marco Argentano (O Cuccurullo), Sarzana (R Petacchi, D Bertoli), Terni(M Bernardinangeli, G Proietti, G Proietti), Trento Villa Bianca (G Cioffi, E Buczkowska),Trento Santa Chiara (P Zeni, C Giovannelli), Trieste ASS 1 (C Mazzone, D Radini), Trieste Università (A Aleksova)

## Footnotes

- **Contributor**s LS, JM and RL had full access to all the data in the study and take responsibility for its integrity and the accuracy of data analysis. Study concept and design: RL and LS. Acquisition, analysis or interpretation of data: LS, JM and RL. Drafting the manuscript: LS, RL and JM. Critical revision of the manuscript for important intellectual content: LS, JM, SM, DN, MD, APM, UHW-T and RL. Statistical analysis: JM.
- **Funding** The GISSI studies and this substudy are supported by the Associazione Nazionale Medici Cardiologi Ospedalieri and by the Istituto di Ricerche Farmacologiche Mario Negri IRCCS. Novartis Pharma Italy funded the GISSI-AF trial. The substudy was partially supported by Roche Diagnostics GmbH Penzberg, Germany. The funders had no role in the design and conduct of the study; data collection, management, analysis and interpretation; preparation, review or approval of the manuscript; and decision to submit the manuscript for publication.
- All authors had full access to all data and take responsibility for the integrity and accuracy of the data analysis. All authors contributed to revision of the manuscript and accepted the final version.
- **Competing interests** SM, LS, APM, MGF, MD and RL received institutional research support from Novartis Pharma and Roche Diagnostics. UHW-T is an employee of Roche Diagnostics GmbH, Penzberg, Germany, the company that funded part of the present study.
- **Patient and public involvement** Patients and/or the public were not involved in the design, or conduct, or reporting, or dissemination plans of this research.
- **Patient consent** Not required.
- **Ethics approval** This study was conducted with the approval of the local ethics committees from the 36 Italian clinical centers that participated in the biomarkers and echocardiographic substudy of the GISSI-AF trial.
- **Provenance and peer review** Not commissioned; externally peer reviewed.
- **Data availability statement** The entire database of GISSI-AF can be accessed upon reasonable request to the Steering Committee.

## Glossary

GISSI-AF trial: Gruppo Italiano per lo Studio della Sopravvivenza nell’Infarto Miocardico–Atrial Fibrillation trial
ARISTOTELE trial: Apixaban for Reduction in Stroke and Other Thromboembolic Events in Atrial Fibrillation

